# CTEPH has shared and distinct genetic associations with pulmonary embolism in a genome-wide association study

**DOI:** 10.1101/2023.05.30.23290666

**Authors:** James Liley, Michael Newnham, Marta Bleda, Katherine Bunclark, William Auger, Joan Albert Barbera, Harm Bogaard, Marion Delcroix, Timothy M. Fernandes, Luke Howard, David Jenkins, Irene Lang, Eckhard Mayer, Chris Rhodes, Michael Simpson, Laura Southgate, Richard Trembath, John Wharton, Martin R Wilkins, Stefan Gräf, Nicholas Morrell, Joanna Pepke Zaba, Mark Toshner

## Abstract

**Background:** Chronic Thromboembolic Pulmonary Hypertension (CTEPH) involves formation and non-resolution of thrombus, dysregulated inflammation, angiogenesis and the development of a small vessel vasculopathy. We aimed to establish the genetic basis of CTEPH to gain insight into these pathophysiological contributors.

**Methods:** We conducted a genome-wide association study (GWAS) on 1945 European cases and 10491 European controls. We co-analysed our results from CTEPH with existing results from GWAS on deep vein thrombosis (DVT), pulmonary embolism (PE) and idiopathic PAH (IPAH).

**Findings:** Our primary GWAS revealed genetic associations at the *ABO*, *FGG*, *TAP2*, *F2*, and *TSPAN15* loci. Through levered analysis with DVT and PE we demonstrate further CTEPH associations at the *F11*, *EDEM2*, *SLC44A2* and *F5* loci but find no statistically significant associations shared with IPAH.

**Interpretation:** CTEPH is a partially heritable polygenic disease, with related though distinct genetic associations to PE and to DVT. The genetic associations at *TAP2* suggest a potential autoimmune component in CTEPH pathology, and the differential effect size of the *F5* association in CTEPH compared to PE/DVT, suggests a lower risk of *F5* polymorphisms in CTEPH.

**Funding:** This study was supported by the NIHR cardiorespiratory BRC and an unrestricted grant from Bayer Pharmaceuticals

**Research in context:** 

**Evidence before this study:** This study is the first genome-wide association study (GWAS) in Chronic Thromboembolic Pulmonary Hypertension (CTEPH). There is some existing evidence for genetic associations in the disease: a European study found an increased CTEPH risk in non-O blood groups and large GWAS have been conducted on CTEPH-related diseases pulmonary embolism (PE) and deep vein thrombosis (DVT). A literature review (MedLine and Google Scholar; 14 Dec 2020) using the keywords ‘Chronic Thomboembolic Pulmonary Hypertensions’ or ‘CTEPH’ and ‘genetic’ showed that no other genetic associations with CTEPH have been reported at genome-wide significance (p < 5 x 10^-8^).

**Added value of this study:** This study reports several new genetic associations with CTEPH, and identifies similarities and differences between the genetic architectures of CTEPH and DVT/PE. Shared and differential genetic associations between CTEPH and DVT/PE may lead to insights into disease pathobiology and help in developing the potential for use of genetic markers in CTEPH risk prediction

**Implications of all the available evidence:** CTEPH is associated with multiple genetic variants that include *ABO*, variants adjacent to the *FGG*, *TAP2*, *TSPAN15*, *F2*, *F5/NME7*, *F11*, *SLC44A2* and *EDEM2* genes. CTEPH has a similar but not identical genetic architecture to PE and to DVT. There is no evidence of shared genetic architecture with idiopathic pulmonary arterial hypertension.

## Introduction

Chronic thromboembolic pulmonary hypertension (CTEPH) is characterised by the organisation and fibrosis of thromboembolic material leading to the obstruction of proximal pulmonary arteries which, together with a secondary small-vessel vasculopathy, results in pulmonary hypertension and subsequent right heart failure.

CTEPH is conventionally considered to result from a process of disordered thrombus resolution following one or more episodes of acute pulmonary embolism (PE) (1). The pathobiology of thrombus non-resolution following acute PE however remains poorly understood but likely arises from complex interactions between mediators of the coagulation cascade, angiogenesis, platelet function and inflammation in association with host factors. Large volume acute PEs, idiopathic presentation, and PE recurrence are associated with a risk for CTEPH development (2). Inefficient anticoagulation may also trigger thrombus formation (3). These factors however do not serve to explain the development of CTEPH in most patients. Furthermore, up to 25% of CTEPH patients do not have a history of antecedent PE. The ability to identify abnormalities in coagulation/fibrinolysis pathways in CTEPH patients is compounded by their treatment with therapeutic anticoagulation and lack of a good animal model of CTEPH.

Genetic studies in CTEPH have the potential to inform our understanding of disease pathophysiology, but have thus far been hampered by the challenge of assembling cases in rare diseases. A European prospective registry found an increased CTEPH risk in non-O blood groups, in a similar pattern to DVT and PE (4), indicating a genetic association with the disease at this locus. This differential risk with ABO is also seen in overall risk of PE and other clotting disorders. To our knowledge, no other genetic associations with CTEPH have been confirmed at genome-wide significance (P< 5 x 10-8).

The genetic basis of a comparator disease, Idiopathic Pulmonary Arterial Hypertension (IPAH) has been much more systematically explored. Heterozygous germline mutations in *BMPR2* are found in 10 – 20% of individuals with IPAH alongside rarer sequence variants including *SMAD9*, *ACVRL1*, *ENG*, *KCNK3* and *TBX4* (5). A more recent GWAS study has also identified common variants contributing to IPAH aetiology and clinical course (6).

An improved understanding of the genetic basis of CTEPH has the potential to not only inform disease aetiopathogenesis but in quantification of CTEPH risk, preventative strategies and treatment options. An evaluation of CTEPH genome-wide associations is therefore warranted. Co-analysis with existing GWAS in PE and DVT aims to improve both discovery and the interpretation of results in comparison to other venous thromboembolic phenotypes. Given well-known genetic drivers to the development of IPAH and its shared pathobiological features of vascular remodelling, inflammation and dysregulated angiogenesis with CTEPH, genetic associations between CTEPH and IPAH were also explored.

## Methods

### Study samples and participants

The study was approved by the regional ethics committee (REC no. 08/H0802/32 and 08/H0304/56). All study participants provided written informed consent from their respective institutions.

We conducted a two-stage design: a discovery study including only UK samples, and a replication stage using non-UK cases and a mixture of non-UK and UK controls.

CTEPH was diagnosed using 2015 ESC/ERS guideline international criteria, and patients were excluded if they had other major contributing factors to their pulmonary hypertension. Demographics of CTEPH samples are reported in Supplementary Table 1. Controls were sourced randomly from the population (without requiring absence of thromboembolic phenotypes). Samples in the discovery phase were genotyped on one of four platforms: the Illumina HumanOmniExpress Exome-8 v1.2 BeadChip (1555 cases, 1693 controls); the Illumina HumanOmniExpressExome-8 v1.6 BeadChip (372 cases, 12 controls); the Affymetrix Axiom Genome-Wide CEU 1 Array (541 cases, 5984 controls, including re-genotyping of 1533 controls genotyped on the Illumina HumanOmniExpressExome-8 v1.2 BeadChip) and the Affymetrix UK Biobank Axiom array (6717 controls).

We performed sample-and SNP-wise quality control on our dataset (7) and excluded cases of non-European ancestry using principal components generated using the 1000 genomes project. We imputed all genotypes to whole-genome cover using the Haplotype Reference Consortium panel on the Sanger imputation server (8,9), separating samples by genotyping platform, and we included SNPs with an INFO score of at least 0.5 across all genotyping platforms used in the study. Full details of quality control procedures are given in the Supplementary Methods.

We separated the discovery cohort into two groups by genotyping platform (Affymetrix or Illumina) and analysed each separately. In each cohort, we used a logistic regression with ten principal component covariates to generate association statistics, and corrected results for residual genomic inflation (10). Since each analysis involved separate samples, we combined results across platforms using a routine p-value meta-analysis using Fisher’s method accounting for effect directions.

We co-analysed our p-values from the CTEPH meta-analysis with p-values derived from a GWAS on self-reported PE drawn from the UK Biobank (11) (GWAS round 2; self-reported DVT (code 20002_1094) and self-reported PE (code 20002_1093)). Details of the co-analysis are given in the supplementary material. In short, the output of each co-analysis is a set of p-values for CTEPH ’adjusted’ for the overall genetic similarity between CTEPH and the second disease (12), which we call ’V-values’. We also performed an analysis using results from DVT in place of results from PE, but found the results from the two analyses were very similar, so we focus principally on the analysis of PE.

Noting that our replication cohort was analysed at genome-wide SNPs, we defined genetic associations at three tiers of significance, all of which generally correspond to a genome-wide significance of overall p-value < 5 x 10^-8^ with varying levels of evidence in the discovery and replication sub-cohorts. The first tier required P < 5 x 10^-6^ in the combined discovery cohort, P < 5 x 10^-3^ in the replication cohort, and P < 5 x 10^-8^ in the combined meta-analysis, with consistent directions of effect across the two sub-analyses in the discovery study and in the replication study. The second tier, designed to ensure nominal association in each cohort and overall genome-wide significance, required a nominal association of P < 5 x 10^-2^ in discovery and replication cohorts and P < 5 x 10^-8^ in the overall meta-analysis, again with consistent directions of effect. The ‘adjusted’ p-values allowed a comparison of evidence for association using cFDR in a similar way to a comparison using meta-analysed p-values, and hence we defined a third tier of association requiring a p-value of 5 x 10^-8^ in either the overall meta-analysis or the ‘adjusted’ sets of p-values derived from leverage of the CTEPH summary statistics on summary statistics for PE, along with consistent directions of effect in discovery and replication cohorts. All p-value thresholds used in ’tier’ definitions were chosen prior to observing the data.

There was a distribution of cases and controls across genotyping batches which could enable confounding batch effects, and differing sources of cases and controls in the replication cohort necessitated across-platform comparisons and imperfect geographical matching resulting in high inflation in association statistics. We also noted recent work indicating that blood-bank sourced control samples may have differing distributions of ABO blood groups to the general population, potentially biasing association statistics at that locus. In the supplementary material, we analyse allele frequencies across batches and cohorts directly, and thus demonstrate that these confounding effects are unlikely to drive our positive associations.

The study design is outlined in figure 1.

**Figure 1.**
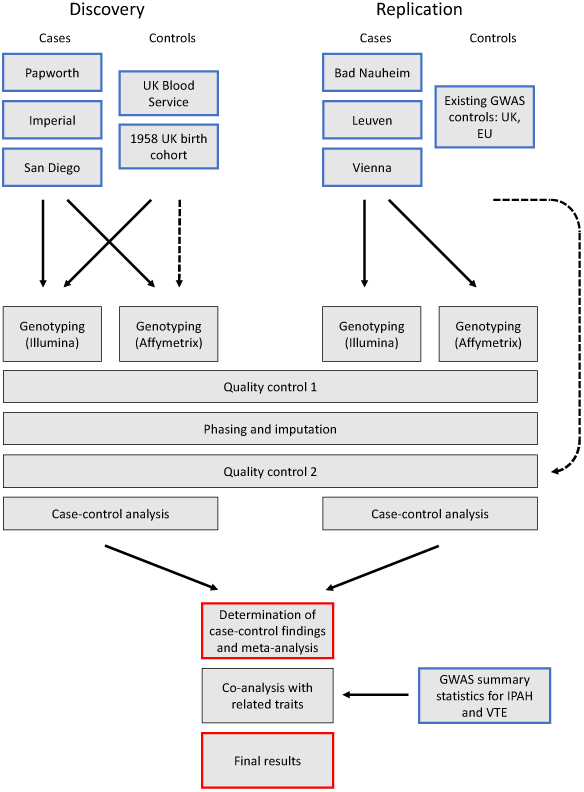
Flow chart for study design.

## Results

### GWAS on CTEPH

After quality control, our dataset consisted of 1146 cases and 5498 controls in the discovery cohort, and 799 cases and 4993 controls in the replication cohort. A total of 4655481 SNPs passed quality control and were included in the final analysis. At tier 2 significance, the study had approximately 80% power to detect an odds ratio of 1.3 for a SNP of minor allele frequency (MAF) 0.25, or an odds ratio of 1.7 for a SNP of MAF 0.05. Further details of power for tier 1 and 2 significance are shown in supplementary figures 1,2,3. Minimal detectable effect sizes at tier 3 significance are more complex; see Supplementary Methods.

Genomic inflation factors (λ) were <1 in the discovery cohort and 1.37 in the replication cohort (λ_1000_ = 1.27 (13)). We were not able to reduce inflation in the replication cohort by inclusion of further covariates or by use of linear mixed models, and concluded that the degree of inflation was inevitable given the imperfect geographical matching between cases and controls in the replication dataset. We corrected P-values in each cohort for this residual inflation (10).

A Manhattan plot of meta-analysed p-values is shown in figure 2, and Table **Error! Reference source not found.** shows peak SNPs in regions reaching genome wide significance. Manhattan plots for the discovery and replication cohorts alone are shown in supplementary figures **Error! Reference source not found.**, **Error! Reference source not found.**. Two regional associations (*FGG* and *ABO*) were found at tier 1 significance, and three further associations (*TAP2*, *TSPAN15*, and *F2*) at tier 2 significance. One further region (*FGG*) reached tier 3 significance on the basis of meta-analysis p-value. Results are shown in table **Error! Reference source not found.**.

**Figure 2:**
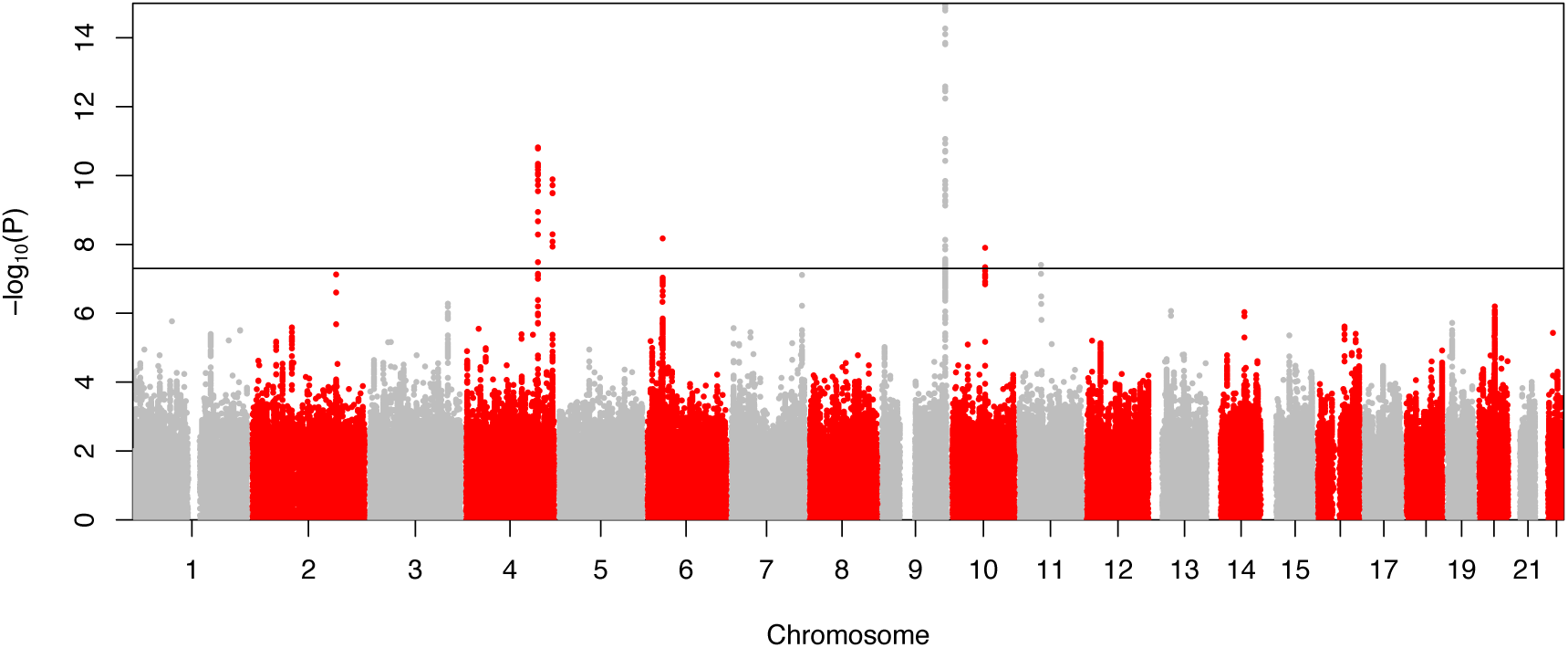
Manhattan plot of p-values derived from meta-analysis of discovery and replication cohorts. The black horizontal line denotes genome-wide significance (*p*=5×10^−8^). The plot is truncated for clarity.

### Co-analysis with DVT and PE

The co-analyses with PE demonstrated three further associations at genome-wide significance (tier 3) Plots of z-scores from the three analysis showed evidence of widespread sharing of associations with DVT and PE, but differential effect sizes between phenotypes (figures 3,4a,4b).

**Figure 3.**
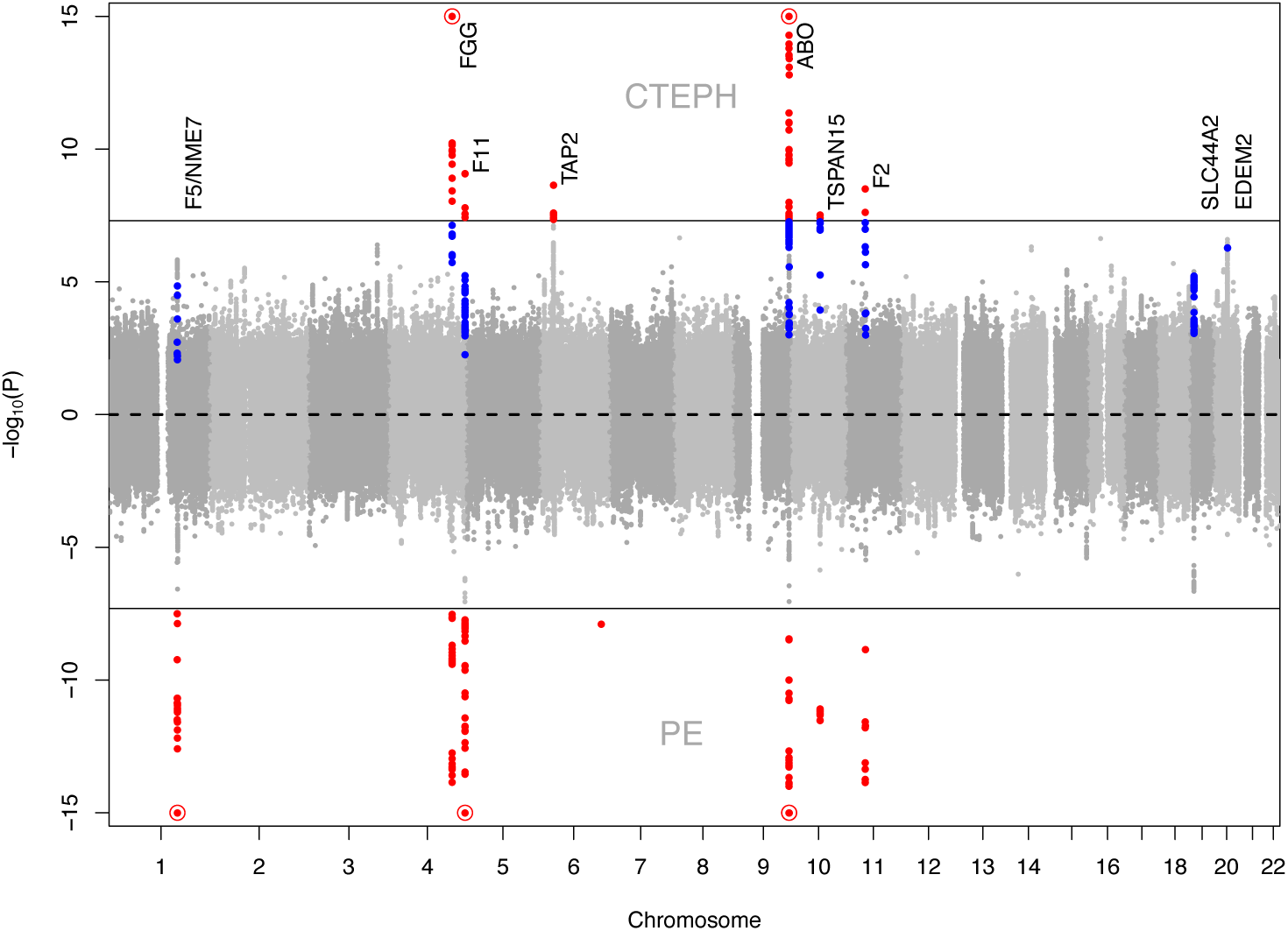
Back-to-back Manhattan plots for CTEPH and PE. The joint associations are clear. Genome-wide associations (p<5 x 10-8) are marked in red. Additional associations discovered through leverage (cFDR) are marked in blue.

**Figure 4a, 4b:**
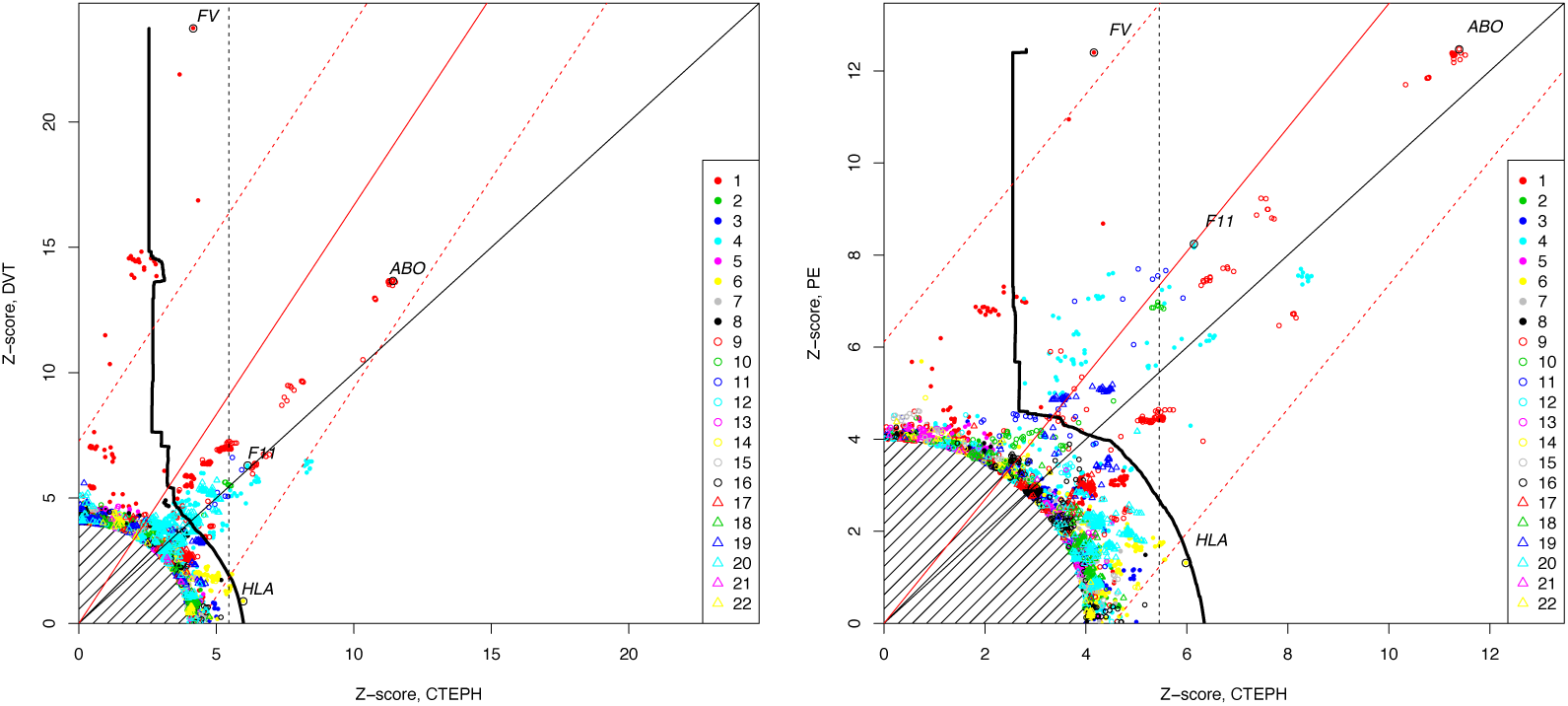
Z-scores for CTEPH against Z-scores for DVT (left) and PE (right). Each point corresponds to a SNP, with colour and shape corresponding to chromosome as per the legend. Z-score pairs close to the origin are excluded. Regions are labelled for some peak SNPs. The area to the right of the dotted black line is a rejection region based on a CTEPH genome-wide significance threshold of *P_CTEPH_* < 5 x 10^-8^. The area to the right of the solid black line is a rejection region based on the levered analysis using conditional false discovery rates, also equivalent to a type-1 error rate of < 5 x 10^-8^. The solid red line shows the expected position of Z-score pairs if SNP effect sizes for CTEPH and DVT/PE were identical. If effect sizes were identical for all SNPs, the probability of any of the points corresponding to the ≈200 SNPs reaching genome-wide significance for CTEPH or DVT/PE falling outside the dashed red lines is <0.05. We see that peak SNPs for *F5* and *TAP2* fall outside the dashed lines in both plots.

### Comparison with IPAH

As a cause of pulmonary arterial hypertension, we considered the possibility that CTEPH shares pathology with idiopathic pulmonary hypertension (IPAH). We did not find genetic evidence of such shared pathology. No shared genome-wide associations are evident between our findings and a recent GWAS on IPAH (6). To assess for genome-scale similarity in genetic basis between IPAH and CTEPH, we used linkage disequilibrium-score regression (LDSC) (14) to estimate genetic correlation *ρ_g_* between the two traits. We also estimated genetic correlation between IPAH and PE (using the summary statistics for PE used in the co-analysis with CTEPH) for comparison.

Diseases with identical genetic bases have genetic correlation 1, and diseases with completely independent genetic bases have genetic correlation 0. If IPAH and CTEPH each occurred as a consequence of some identical underlying cause, we would expect them to have genetic correlation 1, whereas if they were caused by completely independent pathological processes, the genetic correlation would be 0 (and likewise for PE and CTEPH). The observed genetic correlation between IPAH and CTEPH was not significantly different from 0 (est. *ρ_g_* -0.37, standard error 0.38; p-value 0.3 against H^0^: *ρ_g_* =0), but was significantly different from 1. The genetic correlation between CTEPH and self-reported PE was significantly above zero, indicating shared genetic architecture (est. *ρ_g_* 1.07, standard error 0.44; p-value 0.014 against H^0^: *ρ_g_* =0) but not significantly different from 1, indicating that identical genetic architecture could not be ruled out with this analysis. We concluded that, on the basis of genetic correlation, CTEPH is more similar to PE than to IPAH.

### CTEPH GWAS associations

#### FGG and ABO (tier 1)

We found an association with peak SNP rs7659024, around 4kb downstream of the *FGG* gene. The *FGG* gene codes for the gamma chain of the fibrinogen protein, a precursor for fibrin, the principal non-cellular component of blood clots. Polymorphisms in *FGG* are well-known to be associated with DVT (15). The variant is also 9kb downstream of the *FGA* gene, which codes for the alpha chain of the fibrinogen complex. The strongest association (by p-value) in CTEPH was rs687289 in the *ABO* gene, which determines ABO blood group. This locus is also known to be associated with DVT (15). Patients with non-O blood groups are at higher risk of CTEPH (16).

#### TAP2, TSPAN15, F2, SLC44A2, F11 (tier 2/3)

An association was found at tier 2 significance in the *TAP2* gene, which to our knowledge has not been shown to be associated with DVT or PE. SNPs in *TAP2* have been found to be associated with a range of immune-related phenotypes: immunoglobin levels (17), mouth ulcers (18), tuberculosis susceptibility (19), and sarcoidosis (20). Variants rs78677622 and rs149903077 were found at tier 2 significance, and rs2288904 and rs2289252 at tier 3. Variant rs78677622, on chromosome 10, is an intron variant 10kb upstream of *TSPAN15*, which is known to be associated with DVT (15). Variant rs149903077 on chromosome 11 is an intron variant in the *DGKZ* gene, but is likely to correspond to an association of CTEPH with the *F2* gene, from which it is 390kb upstream.

Variant rs2288904 on chromosome 19 is a missense variant in the *SLC44A2* gene, variants in which are associated with DVT (15). Variant rs2289252 on chromosome 4 is an intron in *F11*, which codes for coagulation factor 11, variants in which are DVT-associated (15). Variant rs6060288 on chromosome 20 is an intron in the *EDEM2* gene, variants in which are associated with prothrombin time (21). Finally, we found an association at rs796548658 on chromosome 1 at tier 3 significance. Although the peak variant is an intron in the *NME7* gene, it is likely to represent an association of CTEPH with the *F5* gene, which is strongly associated with DVT (15). This association is notable for the relatively small effect size in CTEPH.

#### Comparison of DVT, PE and CTEPH

We found a substantial difference in observed effect sizes of variants in the *F5* gene between DVT, PE and CTEPH. We also noted that the *TAP2* gene is not known to be associated with DVT or PE. We thus assessed whether the difference in observed effects in *F5* and *TAP2* were larger than expected under a null hypothesis of identical effect sizes (and LD patterns) in CTEPH, DVT and PE.

For both regions, the probability of observing effect sizes at least as different as those seen under the null hypothesis was <0.05, using a Bonferroni correction over all variants reaching genome-wide significance for either disease. This is shown in figure **Error! Reference source not found..**

Considered another way, if the observed odds ratio of the peak SNP for *F5* in DVT [**Error! Reference source not found.**] (or SNPs in close linkage disequilibrium) were equal to the true odds ratio in CTEPH, our study would have had >99% power to detect an association at tier 1 significance. Likewise, if the observed odds ratio for the *TAP2* association found in our study corresponded to the true effect size in DVT, then the study on DVT would have >99% power to detect the association.

We conclude that the effect size of causal variants in *F5* and *TAP2* in CTEPH is different to the effect of those variants in DVT and in PE.

## Discussion

We report the first GWAS in CTEPH, comprising a multinational study on a cohort with sufficient power to find common-variant associations of reasonable size. In general, the associations we find are consistent with a shared genetic associations of venous thromboembolism, although we identify important differences in genetic architecture to PE and DVT. CTEPH is a partially heritable polygenic disease: it does not develop randomly amongst patients with pulmonary emboli, nor is development of CTEPH governed entirely by environmental triggers: if this were the case, all genetic associations for both diseases would have identical size (and variants in *F5* and *TAP2* do not). Historical debate has for decades posited that the similarity in pathophysiology, presence of thrombus in some cases of IPAH and absence of index PE in up to a quarter of cases of CTEPH suggests that CTEPH is not simply the consequence of disordered thrombus fibrinolysis but instead a potential overlap of distal cases of CTEPH and IPAH (22). Our work supports evidence that CTEPH and IPAH are distinct and that despite similar vascular remodelling, inflammation and involvement of dysregulated angiogenesis, the underlying aetiologies are different. This is consistent with work examining CTEPH cohort demographics and phenotypes (23). Genetic associations of underlying susceptibility to vascular remodelling or pulmonary hypertension do not appear to be major drivers of CTEPH in this study.

The smaller effect sizes of variants in *F5* in CTEPH may be an example of index-event bias (24). The large effect of the F5 Leiden variant in causing thromboembolic disease may paradoxically mean that patients with PE carrying a F5 Leiden variant have a *lower* burden of other genetic and environmental risk factors for CTEPH, and are hence less likely to develop CTEPH following PE than those without the variant. This could also account for the apparently smaller relative effect of *F5* variants in PE than in DVT seen in figures **Error! Reference source not found.**.

A downstream and important consideration of our work will be the possibility of careful stratification of “at risk” patients early in the course of their PE treatment for more targeted therapeutic studies looking to prevent the non-resolution. Genetic risk scores can also be posited to enrich for patient subgroups suitable for more extensive antithrombotic clinical trials. The association of *TAP2* for example may suggest that trials of additional anti-inflammatory/immunomodulatory therapies in selected patients may be feasible and warrant consideration. To our knowledge, *TAP2* has not previously been shown to be associated with either DVT or PE. TAP2 variants have been described in a range of immune-related phenotypes, reflecting TAP2’s role in the processing and presentation of Major Histocompatibility Complex molecules [24, 25]. Increased CTEPH risk has long been linked with underlying autoimmune and haematological disorders [4]. In addition, a variety of inflammatory cytokines are elevated in CTEPH and correlate with pulmonary artery inflammatory cell infiltration and CTEPH severity [26]. Our finding of an association of CTEPH with the TAP gene further implicates the role of immune dysregulation in the development of CTEPH.

An important shortcoming of our work is the control population in the replication cohort which is not perfectly geographically matched, resulting in a degree of inflation in summary statistics. This is unavoidable with our current dataset, but reassuringly results are appropriately distributed after rescaling of χ^2^ statistics (supplementary figure **Error! Reference source not found.**) and overall findings are not unexpected.

In summary we provide the first large scale GWAS in this rare disease and we demonstrate for the first time the genetic architecture of a complex condition leveraged against comparator datasets. These analyses establish the primacy of dysregulated thrombosis/fibrinolysis in aetiology and extend our understanding of the possible contribution of additional pathophysiological mechanisms including inflammation. CTEPH did not share any genetic associations with IPAH further confirming that despite significant shared pathophysiology these conditions have divergent aetiology.

## Data Availability

GWAS summary statistics from this article are available upon reasonable request to the authors.

## Acknowledgements

We would like to acknowledge the UK tertiary pulmonary hypertension network and the patients who enabled this work. This work was supported by the UK National Institute for Health Research Cardiorespiratory Biomedical Research Council and an unrestricted grant from Bayer Pharmaceuticals. CJR is supported by BHF Basic Science Research fellowships (FS/15/59/31839 & FS/SBSRF/21/31025) and Academy of Medical Sciences Springboard fellowship (SBF004\1095).

## Contributors statement

Author contributions were as follows (using CRediT taxonomy; http://credit.niso.org/):

JL: Data Curation, Formal analysis, Investigation, Methodology, Software, Visualization, Writing original, Writing review and editing

MN: Data Curation, Formal analysis, Investigation, Methodology, Software, Validation, Visualization, Writing review and editing

WA: Resources JAB: Resources

MB: Data Curation, Investigation, Methodology, Software HB: Resources

MD: Resources TF: Resources

SG: Data Curation, Investigation, Software LH: Resources

DJ: Resources IL: Resources EM: Resources

CR: Data Curation, Investigation, Software MS: Resources

LS: Resources

RT: Conceptualization, Resources, Writing review and editing JW: Resources

MW: Resources

NM: Funding acquisition, Project administration, Supervision

JPZ: Conceptualization, Funding acquisition, Project administration, Resources, Supervision, Visualization

MT: Conceptualization, Formal analysis, Investigation, Methodology, Project administration, Resources, Supervision, Visualization, Writing original, Writing review and editing.

Direct data access and verification were performed by JL and MN.

## Supplementary Methods

### Sample details

We recruited Caucasian CTEPH patients from five European and one United States specialist pulmonary hypertension centres: Bad Nauheim (Kerckhoff Heart and Lung Centre, Bad Nauheim, Germany); Papworth (Royal Papworth Hospital, Cambridge, UK), Imperial (Hammersmith Hospital, Imperial College Healthcare NHS Trust, London, UK), Leuven (KU Leuven - University of Leuven, Leuven, Belgium), San Diego (University of California, San Diego, USA), Vienna (Medical University, Vienna, Austria). CTEPH was diagnosed using international criteria (25) and patients were excluded if they had other major contributing factors to their pulmonary hypertension. Cases were recruited between 2011 and 2017. Centres supplied all available bio-banked samples that had been consented for genomic studies and were suitable for DNA extraction. Clinical details of samples are shown in table 1.

**Table 1:**
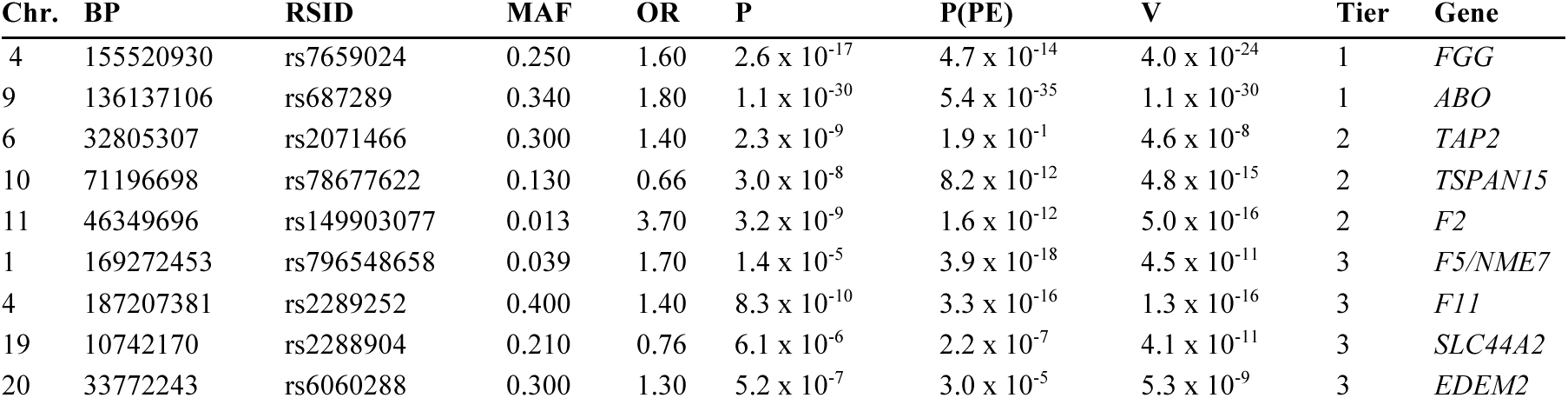
Genome-wide significant regions for CTEPH. Positions shown are GRCh37 and MAF is in controls. Overall odds ratios are estimated from meta-analysis p-values and overall sample sizes. P and P(PE) refer to meta-analysis p-values and p-values for separate GWAS for PE respectively. V are ‘adjusted‘ p-values for CTEPH accounting for overall genetic similarity with PE.

| Chr. | BP | RSID | MAF | OR | P | P(PE) | V | Tier | Gene |
| --- | --- | --- | --- | --- | --- | --- | --- | --- | --- |
| 4 | 155520930 | rs7659024 | 0.250 | 1.60 | $2.6 \times 10^{-17}$ | $4.7 \times 10^{-14}$ | $4.0 \times 10^{-24}$ | 1 | *FGG* |
| 9 | 136137106 | rs687289 | 0.340 | 1.80 | $1.1 \times 10^{-30}$ | $5.4 \times 10^{-35}$ | $1.1 \times 10^{-30}$ | 1 | *ABO* |
| 6 | 32805307 | rs2071466 | 0.300 | 1.40 | $2.3 \times 10^{-9}$ | $1.9 \times 10^{-1}$ | $4.6 \times 10^{-8}$ | 2 | *TAP2* |
| 10 | 71196698 | rs78677622 | 0.130 | 0.66 | $3.0 \times 10^{-8}$ | $8.2 \times 10^{-12}$ | $4.8 \times 10^{-15}$ | 2 | *TSPAN15* |
| 11 | 46349696 | rs149903077 | 0.013 | 3.70 | $3.2 \times 10^{-9}$ | $1.6 \times 10^{-12}$ | $5.0 \times 10^{-16}$ | 2 | *F2* |
| 1 | 169272453 | rs796548658 | 0.039 | 1.70 | $1.4 \times 10^{-5}$ | $3.9 \times 10^{-18}$ | $4.5 \times 10^{-11}$ | 3 | *F5/NME7* |
| 4 | 187207381 | rs2289252 | 0.400 | 1.40 | $8.3 \times 10^{-10}$ | $3.3 \times 10^{-16}$ | $1.3 \times 10^{-16}$ | 3 | *F11* |
| 19 | 10742170 | rs2288904 | 0.210 | 0.76 | $6.1 \times 10^{-6}$ | $2.2 \times 10^{-7}$ | $4.1 \times 10^{-11}$ | 3 | *SLC44A2* |
| 20 | 33772243 | rs6060288 | 0.300 | 1.30 | $5.2 \times 10^{-7}$ | $3.0 \times 10^{-5}$ | $5.3 \times 10^{-9}$ | 3 | *EDEM2* |

In our discovery phase, we compared UK-and California-sourced CTEPH cases to 5984 healthy controls from the UK 1958 birth cohort and UK Blood Service. These samples were originally genotyped on the Affymetrix Axiom Genome-Wide CEU 1 Array, and we re-genotyped 1533 controls on the Illumina HumanOmniExpress Exome-8 v1.2 BeadChip which was used for cases.

In our replication phase, we compared non-UK non-California samples with 6717 UK-and European-samples from a recent GWAS on eosinophilic granulomatosis with polyangiitis (26). Although cases used in the replication dataset were exclusively non-UK, we found that inclusion of UK-sourced controls did not worsen inflation, so we did not restrict control samples to those not from the UK.

### Genotyping, quality control and imputation

As above, our cohort consisted of Illumina-typed cases and controls and Affymetrix-typed cases which we genotyped and imputed, UK-based Affymetrix-typed controls which were previously genotyped, but we imputed, and Affymetrix-typed UK-and Europe-based controls which were previously genotyped and imputed. We were able to split the discovery phase into two separate analyses by platform type, but this was not possible in the replication phase as all controls were genotyped on an Affymetrix platform. Our quality control procedures diverted slightly between the discovery and replication phase.

Illumina samples were genotyped in four separate batches, and Affymetrix cases in a fifth. Genomic DNA was extracted and from whole blood or buffy coat fractions and quantified with ultraviolet-visible spectrophotometry (LGC, Hoddesdon, Herts, UK). DNA was normalised to a concentration of 50*ng*/μ*L* and a total volume greater than 4μ*L* (total DNA >200*ng*), which was required for the DNA microarray. Each batch of micro-array intensity data was normalised and clustered. Genotypes were called independently using Illumina GenomeStudio (v2.0) or the Affymetrix Genotyping Console (4.0). Samples containing more than 1% missing genotypes were removed and SNPs were re-clustered. SNPs with poor clustering quality scores (GenTrain score (<0.7) or clustering separation score (<0.5)) were excluded following re-clustering. Genotyping procedures for the UK 1958 Birth cohort and UK NBS controls chip used in the discovery cohort are described in (27) and for controls used in the replication cohort in (26). We removed samples with heterozygosity rate more than 3 standard deviations from the batch mean or disparate reported and inferred sex. Across all samples including those genotyped, assessed relatedness and removed one of any pair with >30% identity-by-descent, ensuring the absence of first-degree relatives in the dataset.

We then added two further batches: Affymetrix controls from the 1958 birth cohort, and Affymetrix controls from the UK NBS. Within each batch, we removed SNPs with minor allele frequency <1%, SNPs deviating from Hardy-Weinberg equilibrium with p < 1 x 10^-5^, and SNPs with missingness >2% or differential missingness between cases and controls (*p* < 0.05, Bonferroni corrected). We removed samples of divergent ancestry (separating by discovery and replication cohorts), assessed using principal components derived from the 1000 Genomes project (see section below).

We then combined all Illumina samples and all Affymetrix samples into separate combined batches for imputation, and imputed combined batches separately to genome-wide cover (Haplotype Reference Consortium (r1.1)) using the Sanger Imputation Server (8,9), pre-phasing with EAGLE2. Imputation details for replication controls are described in (26). We retained imputed variants with an INFO score of >0.5 and a minor allele frequency of >1% in all three datasets.

We then separated all samples to be used in the replication phase. We combined remaining discovery-phase samples into two cohorts by genotyping platform (Illumina/Affymetrix). Since cases and controls in the replication phase were genotyped and imputed separately, and had somewhat different geographic distributions, we imposed further quality control measures on this cohort. We again removed SNPs with differential missingness between cases and controls (*p* < 0.05, Bonferroni corrected), and removed SNPs with even slightly differing allele frequencies between UK controls in the discovery phase and UK controls in the replication phase (*p* < 0.005).

We then formed three separate cohorts for association testing. We split the discovery cohort by platform (Illumina/Affymetrix) but were unable to do this for the replication cohort, since all control samples were genotyped on an Affymetrix platform, so combined all replication case samples into a single cohort.

### Assessment of divergent ancestry

Principal component analysis using a set of independent directly genotyped SNPs was used to identify samples with outlying ancestry. This was done separately in the discovery cohort (four Illumina batches, combined Affymetrix samples) and with all samples combined in the replication cohort. Samples were initially excluded if they did not cluster with super-populations from 1000 genomes data (28) PCA was then repeated, and samples that did not cluster with 1000 genomes European populations were excluded. Samples were excluded on the basis of distance from the relevant cluster median in standard-deviation units. Thresholds for exclusion were decided visually from each plot, but in no case were samples included if they were more than 3 standard deviations from the median on either the first or second principal component. Plots are shown in supplementary figure 6. Some residual differences can be seen between cases and controls in the replication cohort. Analyses were conducted in R using the snpRelate package (29).

### Statistical analysis

We assessed association between cases and controls using a logistic regression for each cohort, in each case using ten principal components as covariates. Principal components were derived from genotyped SNPs only.

We evaluated genomic inflation in sets of p-values derived from each study. The genomic inflation factor for the replication cohort was large (λ=1.37, λ_1000_ = 1.27) but we were unable to reduce it by inclusion of further covariates or by use of a linear mixed-model (BOLT-LMM (30)) in place of logistic regression. We thus simply corrected for inflation in each cohort by scaling χ^2^ statistics (10).

We combined the two sets of p-values from the discovery cohorts into an overall discovery p-value, and all three sets of p-values into a set of meta-analysed p-values, using a standard z-score meta-analysis. Our criteria for genome-wide association are described in the results overview section above.

### Levered analysis

Define *H^0^_CTEPH_* as a null hypothesis of non-association of a variant of interest with CTEPH. The p-value in our CTEPH GWAS *p_CTEPH_* gives us some information as to whether *H^0^_CTEPH_* holds. We may also be able to glean some information about *H^0^_CTEPH_* holds by considering the association of that variant with some other disease, measured by a p-value *p_OTHER_* from an association study on other on separate samples. This will only be useful if the diseases tend to share the same associations. We use a procedure which both assesses degree of association sharing and tests assocaition with CTEPH in one, involving a quantity termed the conditional false discovery rate, or cFDR (12,31,32). In our case, the ‘other’ phenotype is PE, giving p-values *p_OTHER_* = *p_PE_* (or *p_DVT_*). We consider values (*p_CTEPH_*, *p_PE_*) as samples from the bivariate random variable (*P_CTEPH_*, *P_PE_*) A routine analysis rejection *H^0^_CTEPH_* whenever *p_CTEPH_* < 5 x 10^-8^ corresponds to a rejection subregion of the sample space of the (*P_CTEPH_*, *P_PE_*): specifically, the regions to the right of the dotted black lines in figure **Error! Reference source not found.**. The cFDR replaces this with a data-driven rejection region (the regions to the right of the solid black lines in figure **Error! Reference source not found.**), which approximates the most powerful possible such region (12). It is roughly equivalent to firstly restricting attention to only SNPs for which *P_PE_* < α for some α, concentrating associations with CTEPH.

We can then estimate the joint distribution of p-values for both CTEPH and DVT under the null hypothesis for *H^0^_CTEPH_* and integrate this over these data-driven rejection regions, giving ‘v-values’, which behave like p-values in having uniform distributions under *H^0^_CTEPH_*. These v-values can be thought of as p-values against *H^0^_CTEPH_* ‘adjusted’ for the additional information learned from the set of p-values for DVT association.

### Differential effect sizes between CTEPH, DVT and PE

To determine whether the observed differential effect sizes at *F5* and *TAP2* between CTEPH, DVT and PE reached significance (red lines on figures **Error! Reference source not found.**) we considered a null hypothesis that the underlying odds ratios of these variants were the same in both diseases.

If *n*_1_, *n*_0_, *m*_0_, *m*_1_, μ_1_, μ_0_ represent case/control numbers, observed case/control minor allele frequencies and population case/control minor allele frequencies respectively for a SNP of interest, then the Z score is approximated by

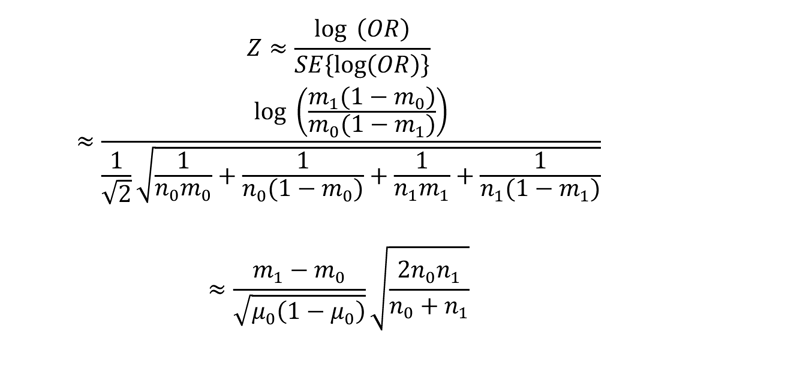

assuming *n_0_* and *n_1_* are large, *μ_0_* ý *μ_1_* and the SNP is diploid. Thus

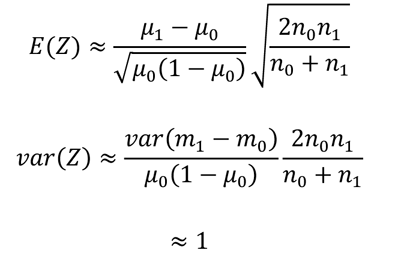

Variance in Z is due to random variance in the study population, and should be independent between studies on independent traits. Thus, denoting *n_1CTEPH_*, *n_0CTEPH_*, *n_1PE_*, *n_0PE_* as case/control numbers in GWAS on CTEPH and PE respectively, under a null hypothesis that the effect size of the SNP is identical in both diseases, the joint distribution of Z scores (*Z_CTEPH_*, *Z_PE_*): will be bivariate normal with mean on a line through the origin with gradient

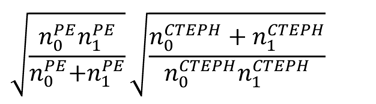

and unit variance *I_2_*. A multivariate normal with unit variance is invariant under rotation, so given *n* SNPs, the probability that at least one pair of Z-scores is at distance greater than *D* from the mean line is approximately

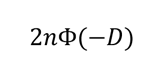

where Φ is the Gaussian CDF function. Dotted lines on plots **Error! Reference source not found.** show distances *D* such that the probability of at least one of the *n* SNPs reaching genome wide significance for either disease lying outside the dotted lines is <0.05.

This is somewhat conservative, since Z scores are dependent due to linkage disequilibrium and the effective number of independent SNPs is less than *n*. Correspondingly, shortcomings of this approach include the possibility that geographic origin can affect relative effect sizes between GWAS and magnitude of linkage disequlibrium between SNPs, potentially confounding the relationship between different disease pathologies and different observed effect sizes in GWAS.

### Power for tier 3 association

To approximate power to reject a variant at tier 3 significance given a z-score *z_other_* at that variant for DVT or PE, refer to the relevant plot in figure **Error! Reference source not found.**. The Z-score that must be obtained for CTEPH in order to reject the null hypothesis for CTEPH equivalent to a p-value < 5 x 10^-8^ corresponds to the x-co-ordinate of the intersection of the horizontal line at *z_other_* with either the dotted or solid black lines (whichever x-intersection gives the smaller value). The minimum odds-ratio resulting in the requisite Z-score given minor allele frequency and study sizes is a straightforward exercise and can be computed from eg. equation **Error! Reference source not found.**.

## Assessment of batch effects

Samples were genotyped in several separate procedures (batches), and between-batch differences (batch effects) could have led to false positive results. The distribution of cases and controls across batches is shown in table 2. The absence of both control and case samples in some batches meant that such batch effects could not be directly differentiated from true case/control differences, and that batch numbers could not be included as covariates in the GWAS analysis.

**Table 2:** Clinical characteristics of case samples, format mean (SD) where appropriate. MPAP: mean pulmonary artery pressure; PVR: pulmonary vascular resistance; CI: cardiac index. Clinical data were not available for all samples.

|  | Pre-QC | Final |
| --- | --- | --- |
| Male (%) | 49.6 | 49.6 |
| Age | 62.5 (15) | 63 (14) |
| Height (cm) | 173 (9.7) | 172 (9.9) |
| Weight (kg) | 82.7 (17) | 84.2 (19) |
| MPAP (mmHg) | 43.3 (13) | 44.2 (12) |
| PVR (dyn . s . cm<sup>-5</sup>) | 679 (400) | 687 (390) |
| CI | 2.45 (0.66) | 2.48 (0.66) |

The three areas of concern were 1. that in the Illumina-genotyped part of the discovery phase, batches 2-4 contained only cases; 2. that in the Affymetrix-genotyped part of the discovery phase, cases and controls were genotyped in separate batches; 3. that in the replication phase, cases and controls were genotyped in separate batches; and 4. that controls in the discovery phase were partially sourced from blood bank samples, which may drive the ABO association through differential distribution of ABO groups.

We address these problems by showing that at our discovered associations, allele frequencies are generally consistent across batches, allowing for case-control status. We also demonstrate that on a genome-wide scale, inter-batch effects are not detectable for each analysis. We acknowledge that the presence of batch effects cannot be definitively ruled out, particularly for the Affymetrix-genotyped part of the discovery phase and for the replication phase.

### Allele frequency at genome-wide associations

We computed allele frequencies across each batch for each genome-wide association in table 3, separating by case/control status. Across these nine associations allele frequencies in batches were generally consistent (Supplementary figure 7).

**Table 3.**
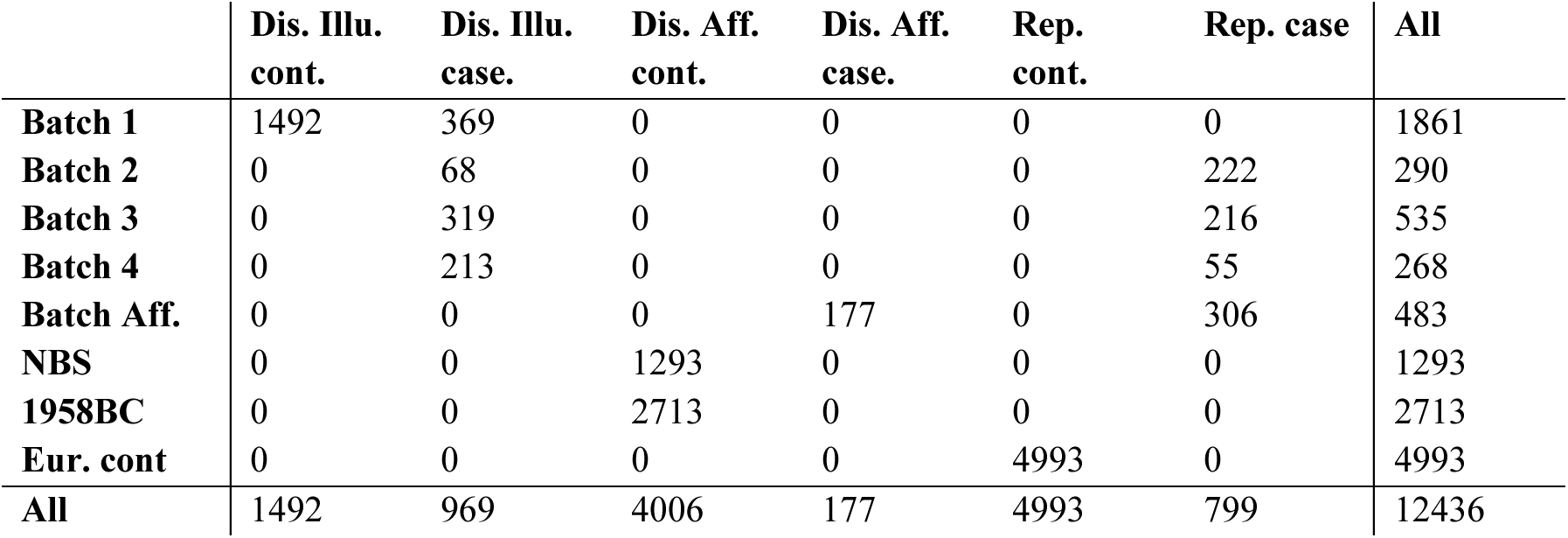
Distribution across batches for cases and controls in the discovery and replication phases. Batches 1-4 used Illumina chips; all other batches used Affymetrix. Cross-platform analyses were not performed in the discovery phase, but were necessary in the replication phase. Genotyping of the final three batches was performed by external groups. cont = control, rep = replication

We also note that the association at the ABO locus (chromosome 9) is not driven by the blood bank-sourced cohort (NBS); allele frequencies for the peak variant are consistent in the NBS cohort and 1958BC cohort, the latter of which, as a birth cohort, can be considered an unbiased population sample. Indeed, allele frequencies are consistent for the NBS and 1958BC cohort for all associations.

### Between-batch comparisons

Where possible, we analysed whether allele frequencies differed systematically across batches within one of the three case/control comparisons. We compared allele frequencies amongst cases in batches 1-4 for the Illumina-genotyped discovery phase, and amongst NBS and 1958BC controls in the Affymetrix-genotyped discovery phase (Supplementary figure 8). Since cases had variable geographic distributions across batches in the replication phase, we could not check this in the same way.

We compared allele frequencies at all variants using Fisher’s exact test, and assessed whether the distribution of resultant p-values differed from the expected distribution of p-values should the observed batches represent identically-genotyped truly random samples from a common population. We found that our results were consistent with this distribution, indicating that systematic batch effects were unlikely to be present.

### Role of the funding sources

Funders had no role in study design; in the collection, analysis, and interpretation of data; in the writing of the report; or in the decision to submit this paper for publication

## STREGA guidelines statement

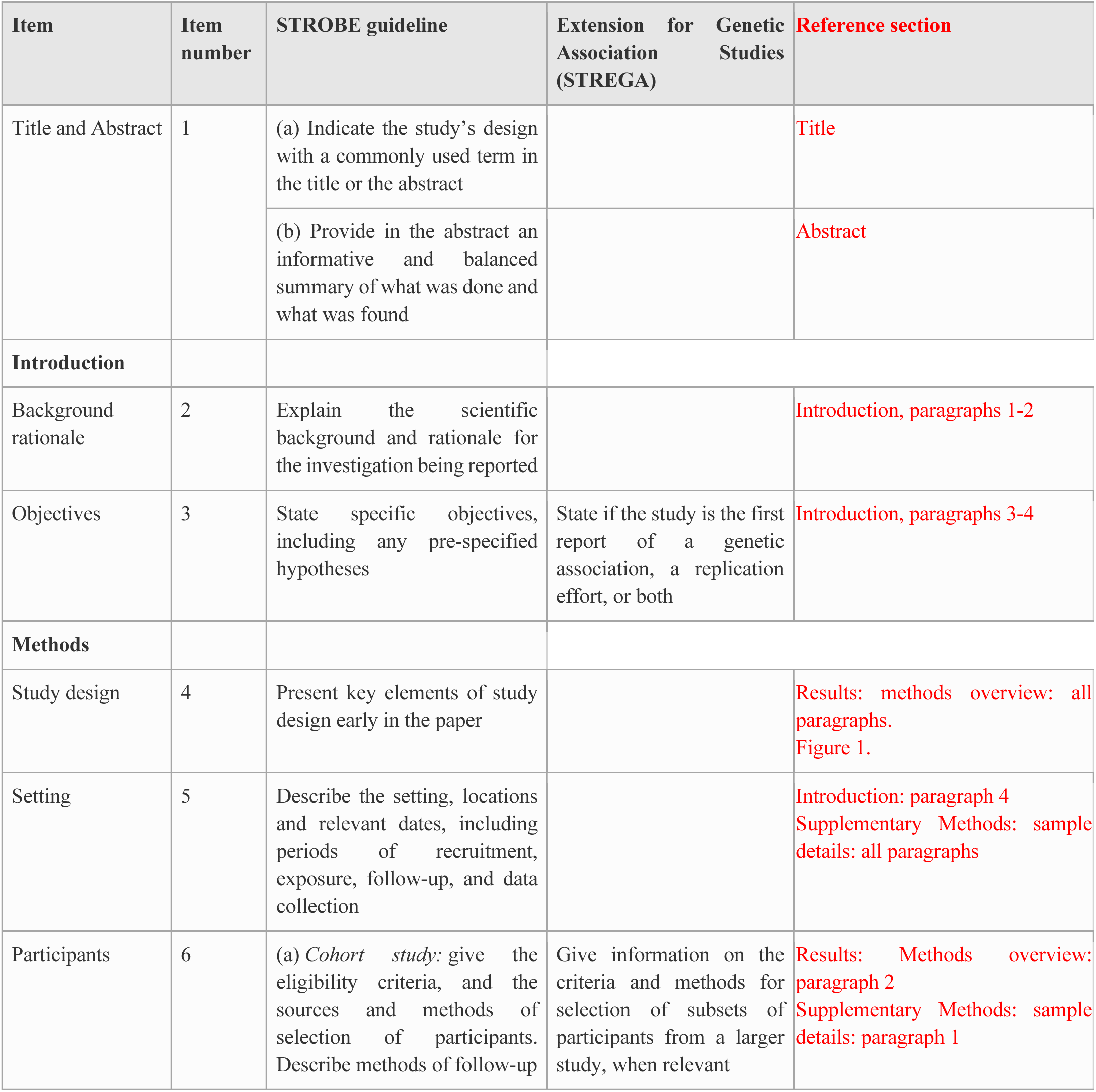
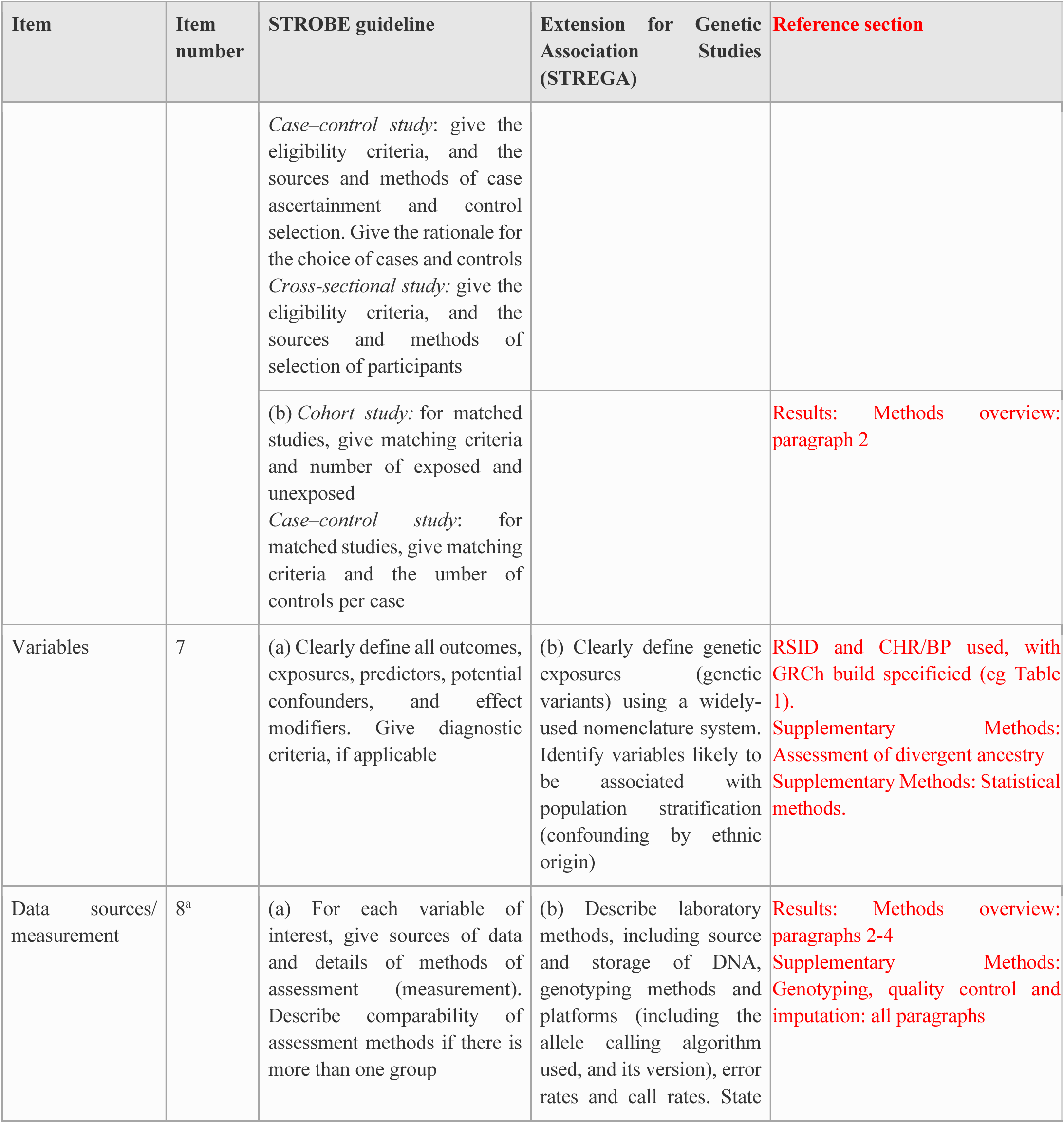
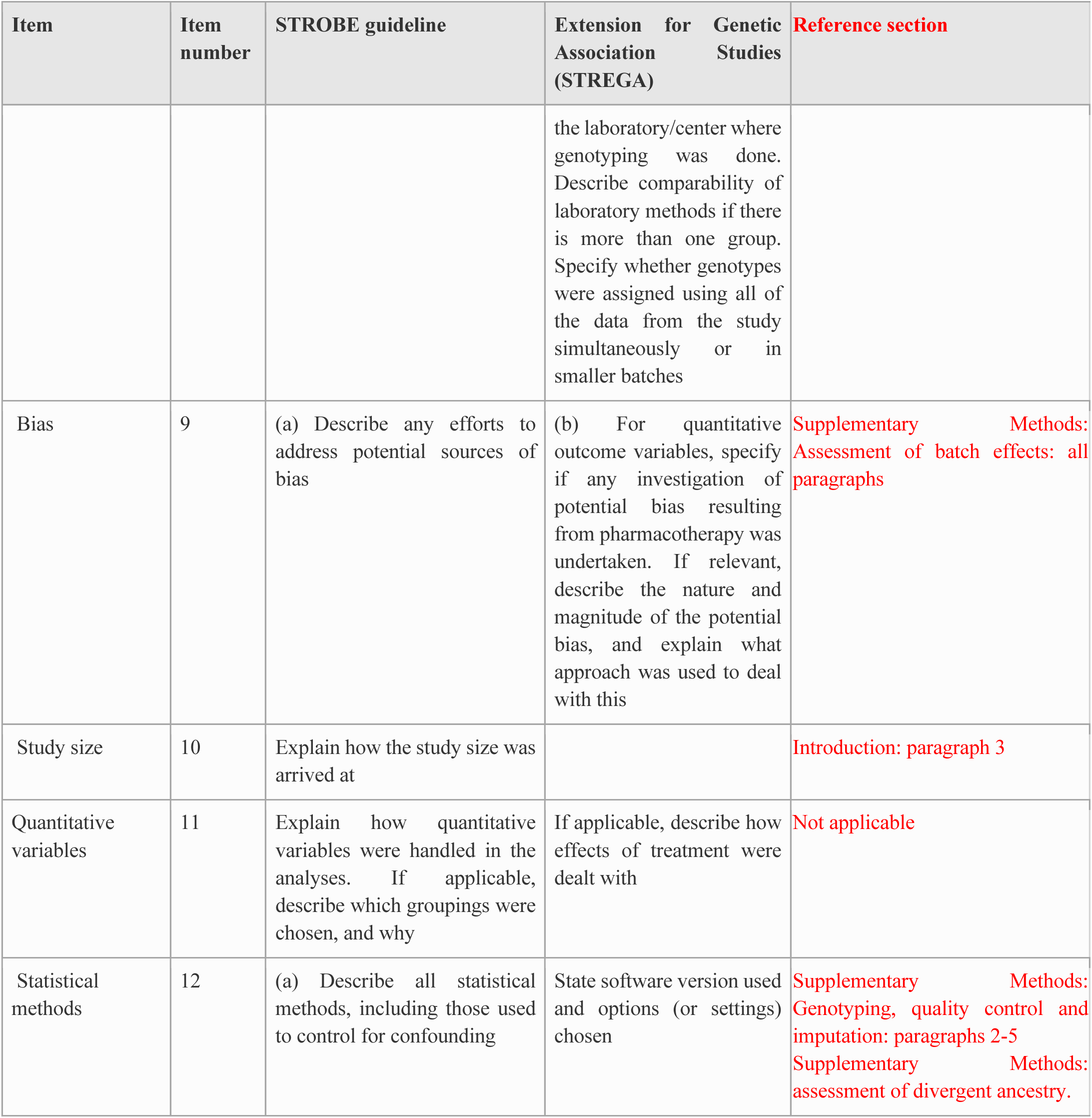
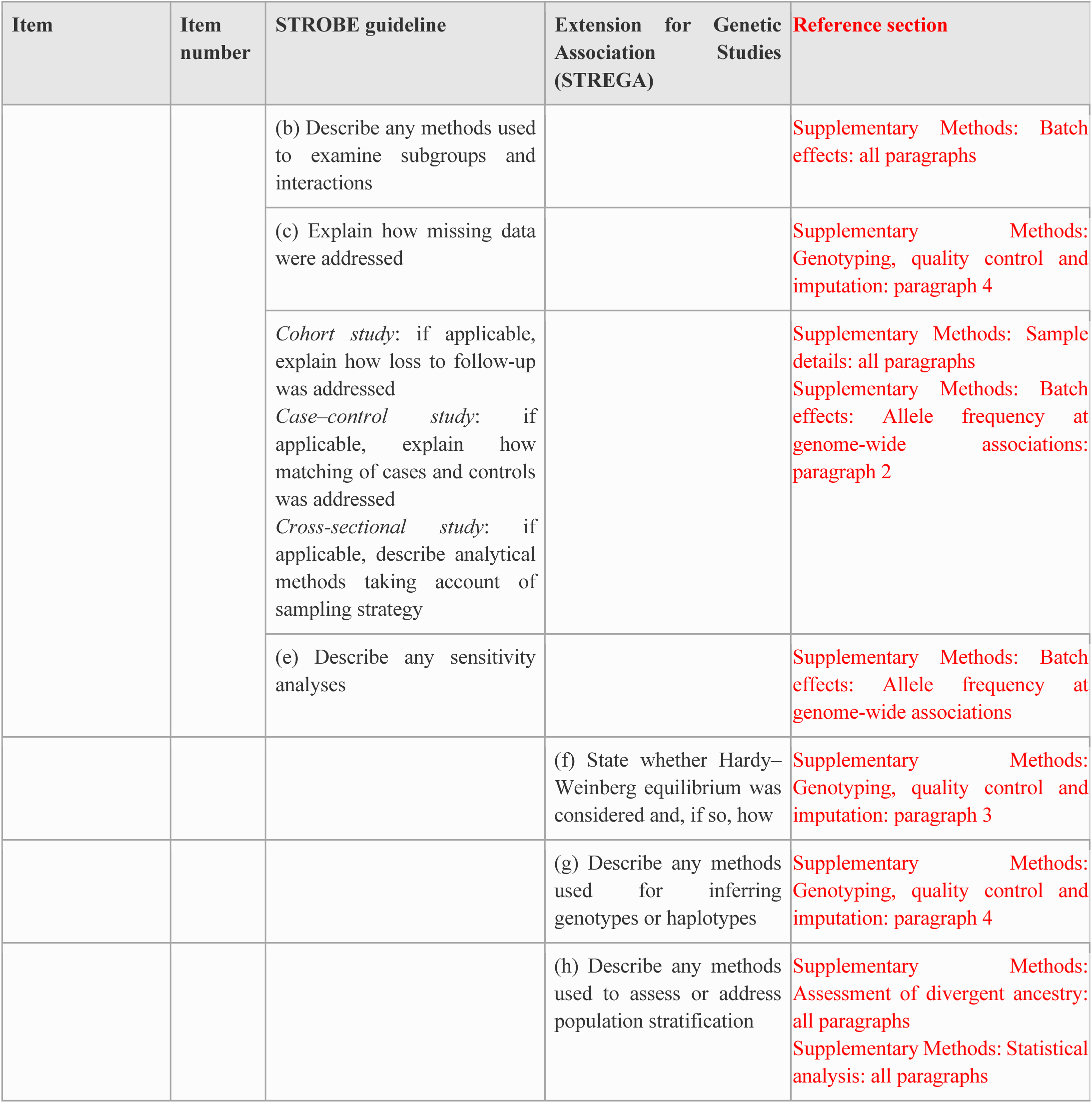
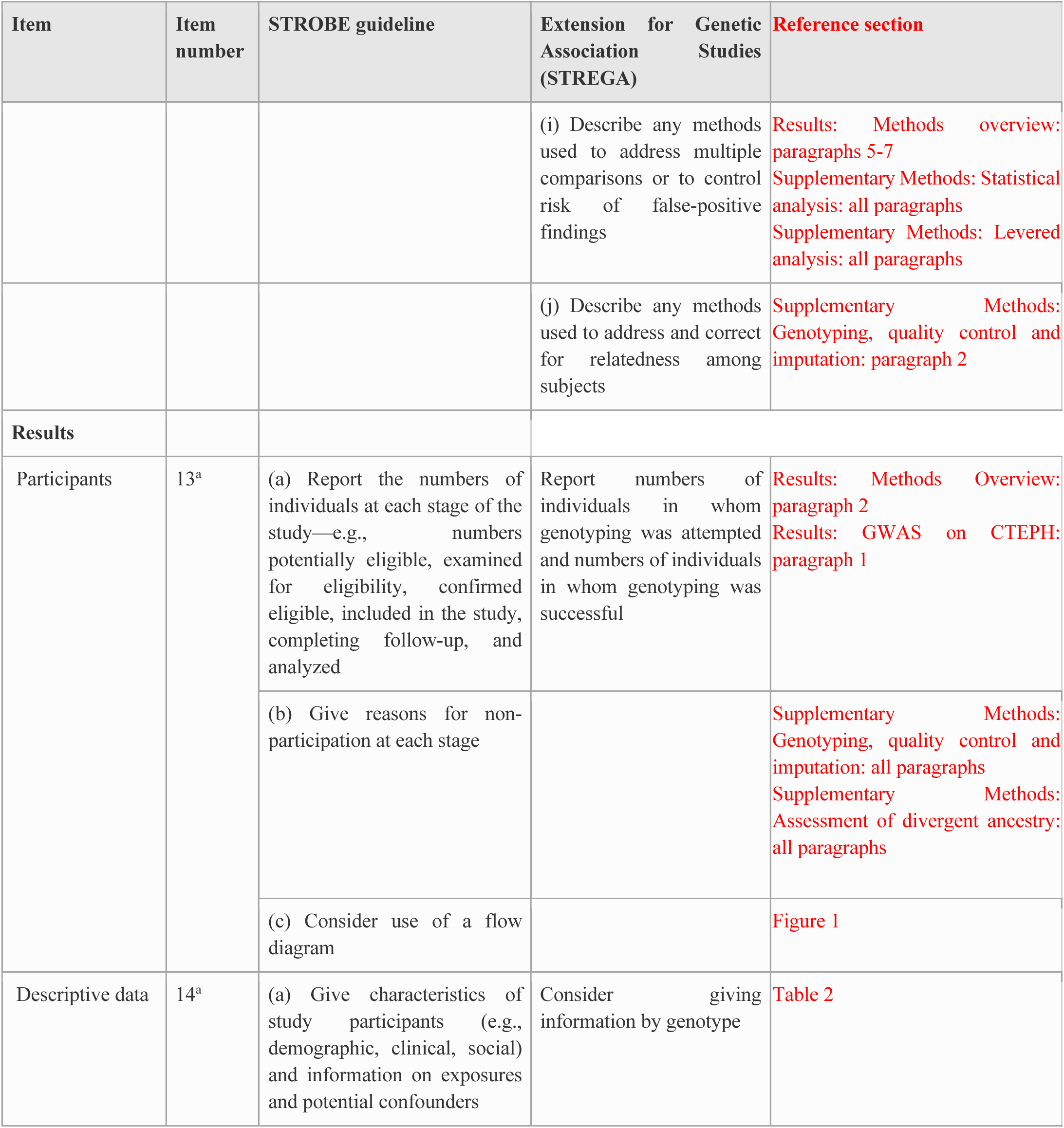
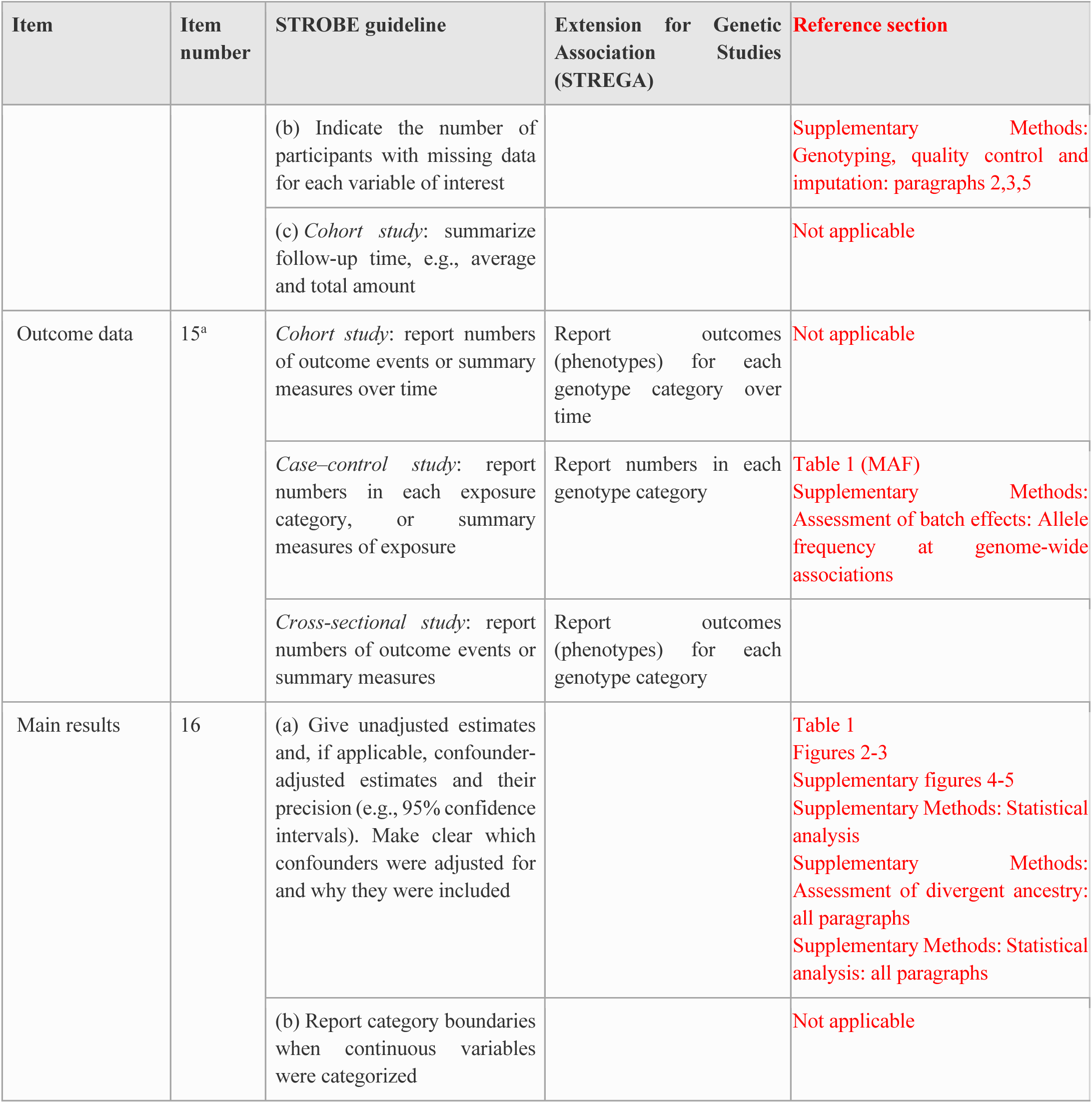
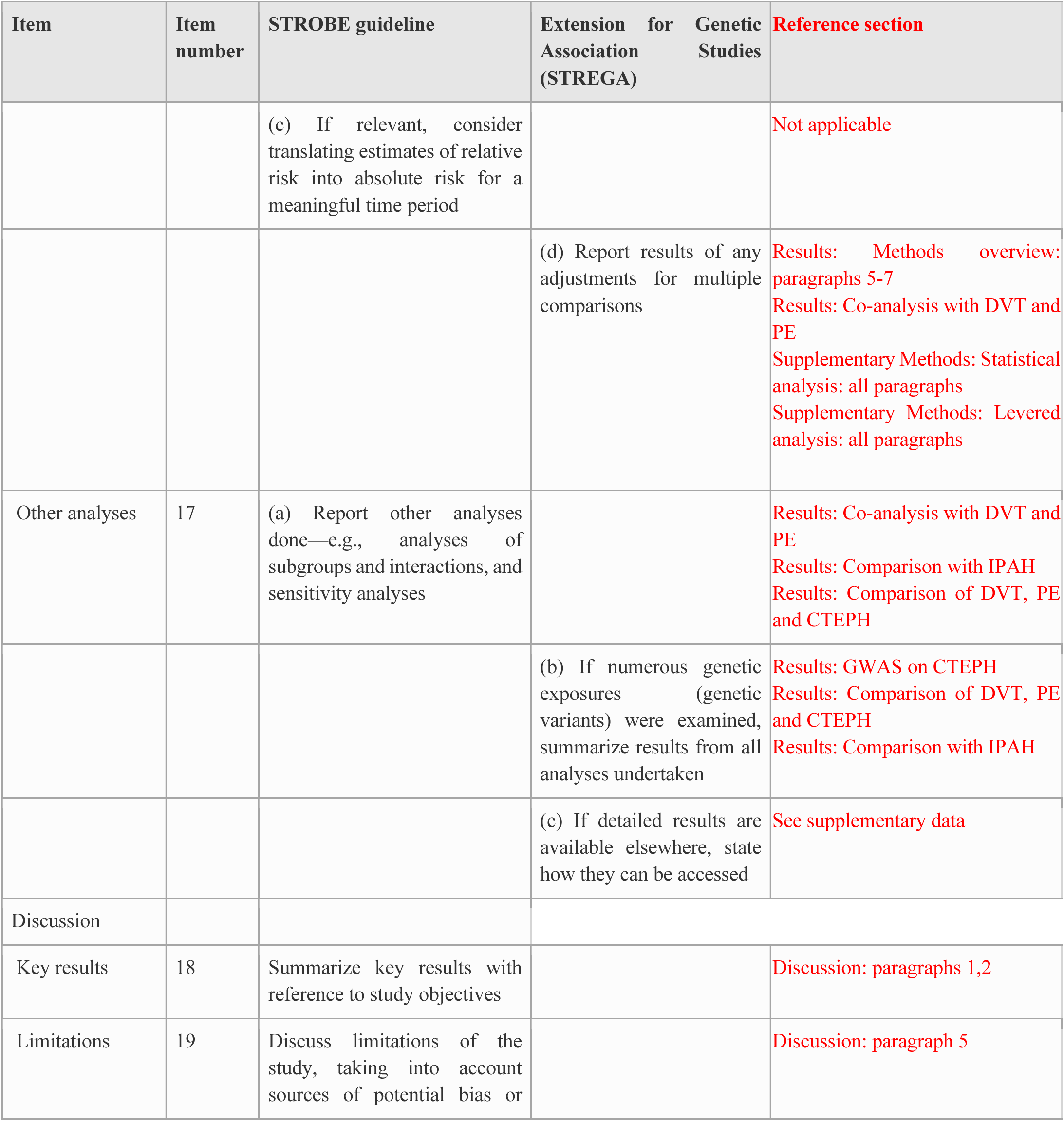
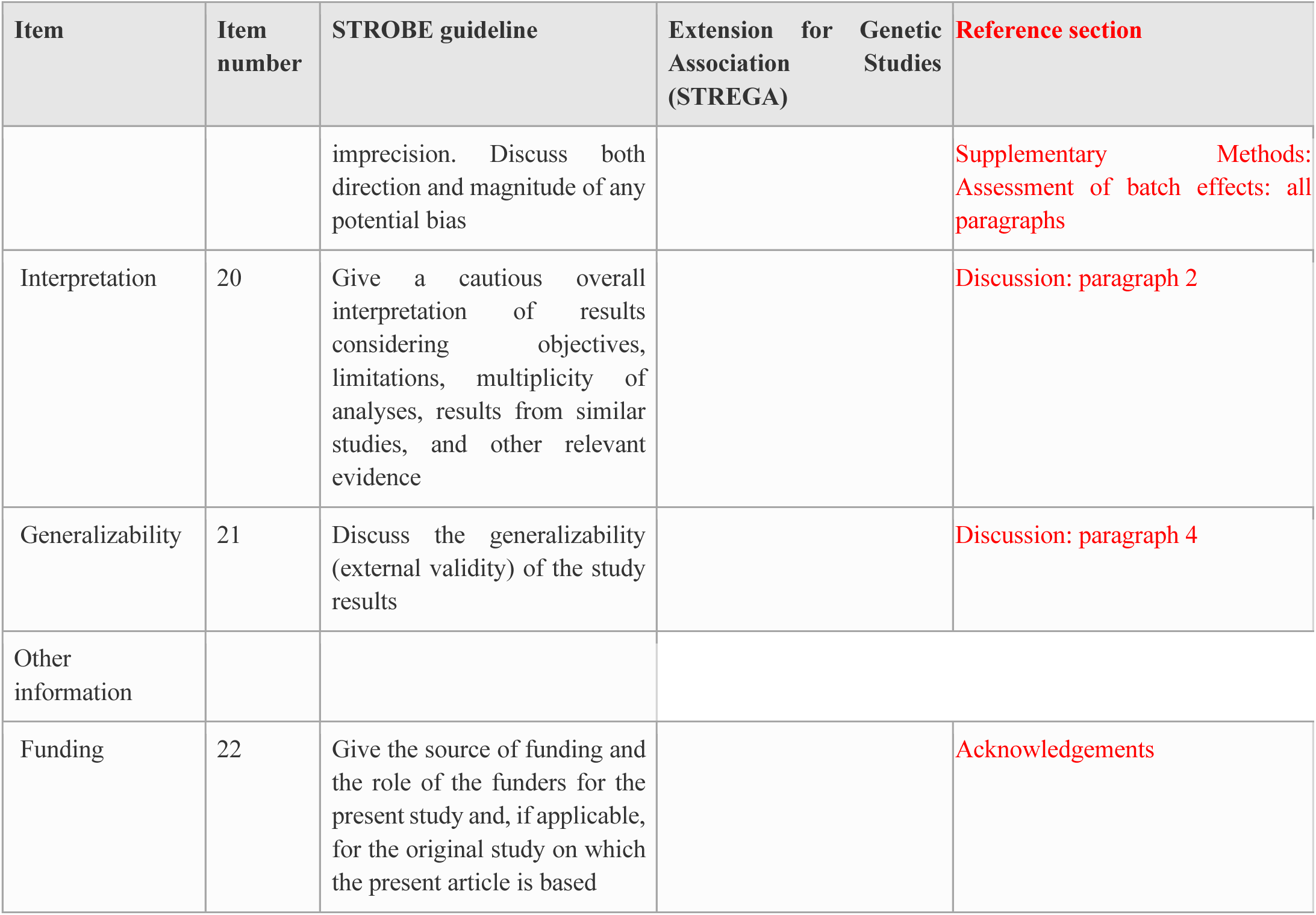

### Supplementary figures

Supplementary figure 1: Power to reject a null hypothesis of CTEPH non-association at tier 1 or 2 significance for a range of of minor allele frequencies in controls. Power calculations take account of the meta-analytic structure but assume no attenuation of power from inclusion of principal component covariates.

Supplementary figure 2: Power to reject a null hypothesis of CTEPH non-association at tier 1 significance for a range of minor allele frequencies in controls and odds ratios.

Power calculations take account of the meta-analytic structure but assume no attenuation of power from inclusion of principal component covariates.

Supplementary figure 3: Power to reject a null hypothesis of CTEPH non-association at tier 2 significance for a range of minor allele frequencies in controls and odds ratios.

Power calculations take account of the meta-analytic structure but assume no attenuation of power from inclusion of principal component covariates.

Supplementary figure 4: Manhattan plot of p-values from discovery cohort. The black horizontal line denotes genome-wide significance (p=5 x 10^-8^)

Supplementary figure 5: Manhattan plot of p-values from replication cohort. The black horizontal line denotes genome-wide significance (p=5 x 10^-8^)

Supplementary figure 6: Principal components of genetic samples combined with 1000 Genomes (1KG) samples. Leftmost plots show principal components including all 1KG samples, middle plots including all European 1KG samples, and rightmost plots including all European 1KG samples after exclusions. Black lines indicate exclusion boundaries.

Supplementary figure 7. Allele frequencies across batches. Horizontal lines show average allele frequencies, and vertical lines show 95% confidence intervals. Occasional inconsistencies for replication subcohorts are likely due to differing geographical distributions.

Supplementary figure 8. Q-Q plot for genome-wide p-values for between-batch comparisons. Confidence intervals are 95% pointwise. Batch differences are consistent with an absence of batch effects.

## Notes

### Competing Interest Statement

CR reports grants, personal fees for lectures, steering committee and advisory board work, and non-financial support for meeting attendance from Actelion, personal fees for lectures and advisory board work from Bayer and MSD, personal fees for advisory board work from GSK, outside the submitted work.
MW reports personal fees from GSK and Johnson & Johnson/Actelion, outside the submitted work.
JPZ received modest fees for research grant, support to travel to meetings and honoraria from Actelion, Bayer, GlaxoSmithKline and Merck Sharp & Dohme
MT reports personal fees from GSK, grants and personal fees from J&J/Actelion, grants from Merck and Bayer, outside the submitted work.

### Author Declarations

UK National Health Service Research Ethics Committee of Cambridge East gave ethical approval for this work (REC no. 08/H0802/32 and 08/H0304/56).

### Summary of Updates

An author (KB) was accidentally omitted from the previous author list. This version corrects that omission.

